# CC- chemokine ligand 25 mediates inflammatory crosstalk between adipose and liver in non-alcoholic fatty liver disease

**DOI:** 10.1101/2021.07.26.21261056

**Authors:** Richard Parker, J Drake, CJ Weston, DH Adams

## Abstract

**Background and Aims:** Obesity is closely associated with non-alcoholic fatty liver disease (NAFLD). We investigated expression of the CC-chemokine CCL25 in adipose tissue (AT) and its relation to hepatic inflammation.

**Methods:** Primary human tissue was used for all experiments. CCL25 concentration in serum was measured with enzyme-linked immunosorbent assay (ELISA). Gene expression of CCL25 was measured with polymerase chain reaction. Protein expression was assessed with ELISA and immunohistochemistry. Leukocyte trafficking was investigated in a dynamic assay to model the hepatic sinusoid. Expression of CCR9 on liver-infiltrating leukocytes analysed with flow cytometry.

**Results:** Circulating CCL25 was increased in obesity. Gene expression of *CCL25* in AT was several-fold higher than in liver, although protein levels of CCL25 were comparable. Soluble CCL25 in flow media increased leukocyte trafficking across hepatic endothelium. Greater numbers of CCR9^+^ cells were seen in liver tissue from patients with NAFLD, where the greatest difference was in the number of CD14^+^CD16^+^ monocytes.

**Conclusions:** CCL25 and its cognate receptor CCR9 mediate a novel pathway of inflammatory crosstalk between adipose and liver tissue in NAFLD.

---

Non-alcoholic fatty liver disease (NAFLD) describes a spectrum of disease from non-alcoholic fatty liver (NAFL) through increasing inflammation (non-alcoholic steatohepatitis, NASH) to cirrhosis and HCC ^[1]^. Since NAFLD was first recognized in the early 1980’s its prevalence has increased dramatically and NAFLD is now the fastest growing indication for liver transplantation in the USA ^[2]^. NAFLD is closely associated with the metabolic syndrome, particularly obesity, where derangements in adipose tissue (AT) function contribute to systemic and hepatic inflammation ^[3]^. Multiple mechanisms underpin the relationship between AT and liver, including lipotoxicity ^[4]^, cytokine release ^[5]^ and alterations in adipokine secretion ^[6]^.

Chemokines are small molecular messengers of diverse function with a canonical role in leukocyte trafficking ^[7]^. These proteins are attractive targets for therapeutic targeting and the last decade has seen development of chemokine antagonists in diverse clinical applications including liver and metabolic diseases such as in obesity where alterations in the expression of chemokines in adipose tissue correlate with the severity of NAFLD ^[8; 9]^. CC-chemokine ligand 25 (CCL25) was first described as playing a role in T lymphocyte development in the thymus while more recent reports have implicated CCL25 in various inflammatory conditions including colitis and associated liver disease ^[10]^. We now report data that describe a role for CCL25 as a mediator of inflammatory crosstalk between adipose and liver tissue in human fatty liver disease.

## Materials and methods

### Human tissue for research

Blood obtained through the haemochromatosis venesection programme at University Hospitals Birmingham was used as a source of leukocytes. All patients gave informed consent and research ethics committee approval was given (reference 06/Q2708/11). Serum for ELISA was collected from healthy volunteers.

Human adipose tissue was collected from patients undergoing abdominal surgery at University Hospitals Birmingham, by the human biomaterials resource centre (HBRC) at the University of Birmingham. All patients gave informed consent for use of samples for research. The HBRC is licensed under the UK Human Tissue act (HTA) (licence 12358). It has research ethics committee approval to collect and distribute samples (reference 15/NW/0079). Adipose tissue was snap frozen or placed in formalin at the time of collection.

Human liver tissue was collected at University Hospitals Birmingham NHS Foundation Trust (UHB). All liver tissue was obtained through the hepatobiliary surgical programme where research Ethics Committee approval has been granted for the consensual use of resected or explanted liver tissue (reference 06/Q2708/11). Liver tissue was snap frozen or placed in formalin at the time of collection.

### Isolation and culture of non-parenchymal cells

Human sinusoidal endothelial cells (HSEC) were isolated from explanted human livers. Liver was finely chopped before being digested with collagenase and strained through sterile mesh. The resulting homogenate, after washing, was layered over 33% and 77% percoll gradients and centrifuged at 2000rpm for 25 minutes. The band of cells at the interface of the 33% and 77% percoll was removed and washed. HSEC were separated by magnetic bead separation using antibodies to human embryonic antigen (HEA) to separate HSEC.

### Immunohistochemistry

#### Paraffin embedded sections

Adipose or liver tissue was placed in formalin until use. Tissue was then embedded in paraffin, cut into 8μm sections and mounted onto glass slides.

#### Immunohistochemistry procedure

Paraffin embedded sections were rehydrated by immersion in clearene (6 minutes) and ethanol (4 minutes). Endogenous peroxidase activity was quenched by immersion in hydrogen peroxidase. Antigen retrieval was performed by incubating slides in high pH antigen retrieval solution (Vector) at 65°C overnight (approximately 16 hours). Sections were washed in TBS, and non-specific binding blocked with casein and horse serum as above. After twice washing in TBS for 5 minutes, primary antibody against CCL25 (diluted to 4μg/mL) or isotype control (rabbit IgG) were added and slides incubated at 4°c for 12 hours. After washing slides three times in TBS, secondary antibodies (IMMpress kits, Vector Labs, USA) were applied for 1 hour at room temperature. NovaRed was applied for 2 minutes before washing in tap water and counterstaining in filtered Mayer’s haematoxylin for 30 seconds. Slides were dehydrated through alcohol (4 minutes incubation) and clearene (6 minutes incubation), and mounted with DPX mountant. Sections were visualised on an Axioskop 40 microscope.

### Protein lysates

Protein tissue lysates were made from adipose and liver tissue. Tissue was homogenized in 20μl of Cellytic MT lysis reagent (Sigma-Aldrich, Missouri, USA) per mg of liver tissue, supplemented with protease inhibitors (Complete Mini, Roche Diagnostics, Indiana, USA). After agitation for 1 hour at 4 °C samples were centrifuged at top speed in a microfuge for ten minutes and supernatant collected. Concentration of protein was measured using BCA reaction (Sigma Aldrich, Missouri, USA) with bovine serum albumin (Sigma Aldrich, Missouri USA) as standard, and sample protein concentration calculated from the standard curve. Protein lysates were stored at −20°c at 500mg/ml until use.

### Polymerase Chain Reaction

#### Isolation of RNA from tissue

RNA was isolated by homogenizing adipose or liver tissue in Trizol (Life Technologies, California, USA). Chloroform was added and samples centrifuged at top speed in a microfuge for 15 minutes. The upper aqueous layer was removed and isopropanol added. Samples were centrifuged at 12,000rpm for 15 minutes. The resulting RNA pellet was washed in 70% ethanol and re-suspended in nuclease free water. Concentration of RNA was measured with a Nanophotometer (Implen, Germany) three times and a mean reading recorded. Purity was noted with 260/280 nm ratio.

#### Synthesis of cDNA

cDNA was prepared from RNA using Taqman reagents (Life Technologies, California, USA) according to the manufacturer’s instructions: briefly, 2 μL of RNA was combined with random hexamers, reverse transcriptase, RNase inhibitor, magnesium chloride and a buffer solution. This mixture was heated to 25°C for 10 minutes, 37°C for 30 minutes, 95°C for 5 minutes and then cooled to 4°C. cDNA was then stored at −20°C until use.

Probe/primer mixes for CCL25 and 18S were obtained from Taqman (Life Technologies, California, USA, catalogue numbers Hs00608373_m1 and Hs03003631_g1 respectively) and made up with Taqman reagents. A 96 well plate was used for reactions, with wells containing cDNA, primer/probe mix (where FAM was used for the probe)(**table 2.5**) and Taqman mastermix. Three replicates were used for the gene of interest and three for housekeeping gene. A water control was used in each experiment, where water was used in place of cDNA. In addition, a reverse-transcriptase control was used, where cDNA synthesis was performed without reverse transcriptase and the resulting, presumed cDNA free, control used to rule out the presence of genomic cDNA in the samples.

PCR experiments were performed using a Roche Lightcycler 480 machine with cycling parameters: 2 minutes at 50°C, 10 minutes at 95oC, followed by 40 cycles of 10 seconds at 95°C and 1 minute at 60°C. A single quantification measurement was taken during each cycle.

### Enzyme Linked Immunosorbent Assays (ELISA)

To measure concentration of CCL25 in serum, a DuoSet ELISA kit from R&D Systems (Cat. No. DY334, R&D systems, Minnesota, US) was used. Whole blood was collected into Vacutainers containing citrate and centrifuged at 2000rpm for 10 minutes to separate serum, within an hour of collection. Serum was collected and stored at −80°C until use. ELISA was performed in accordance with manufacturer’s instructions. Recombinant human CCL25 was used as a positive control (Peprotech, New Jersey USA). Samples were diluted in sample buffer 1:4 and run in duplicate. An ELISA kit from Cusabio (catalogue number CSB-E09189h) was used to measure concentration of CCL25 in protein lysates from whole adipose or liver tissue. In each case a standard curve was generated from known concentrations of recombinant chemokine and sample values interpolated from this curve.

### Flow Cytometry analysis of leukocytes

Liver tissue was dissociated mechanically using a GentleMACS and C-tubes (using program Spleen_01_01; Miltenyi, Germany) and the resulting homogenate passed through a fine mesh (70 micron pores). After washing three -five times with PBS (until a clear supernatant was achieved), 30mL of liver tissue homogenate suspended in PBS was layered over 20mL of Lympholyte (CedarLane Labs, Canada), and centrifuged at 2000rpm/870g for 25 minutes. The resulting mononuclear cell layer was removed and washed a further three times in PBS. Cells were analysed immediately

Isolated leukocytes from liver tissue were suspended in 100μL at 1×10^6^ cells/ml in MACS buffer (PBS containing 2% FCS and 1mM EDTA) and incubated with antibodies as shown in **table 2.6**. After incubation for 20 minutes at room temperature, cells were washed, re-suspended in PBS and analysed by flow cytometry using a Beckman Coulter Cyan. Cells stained with single colours were analysed for compensation and isotype controls were used to define negative populations.

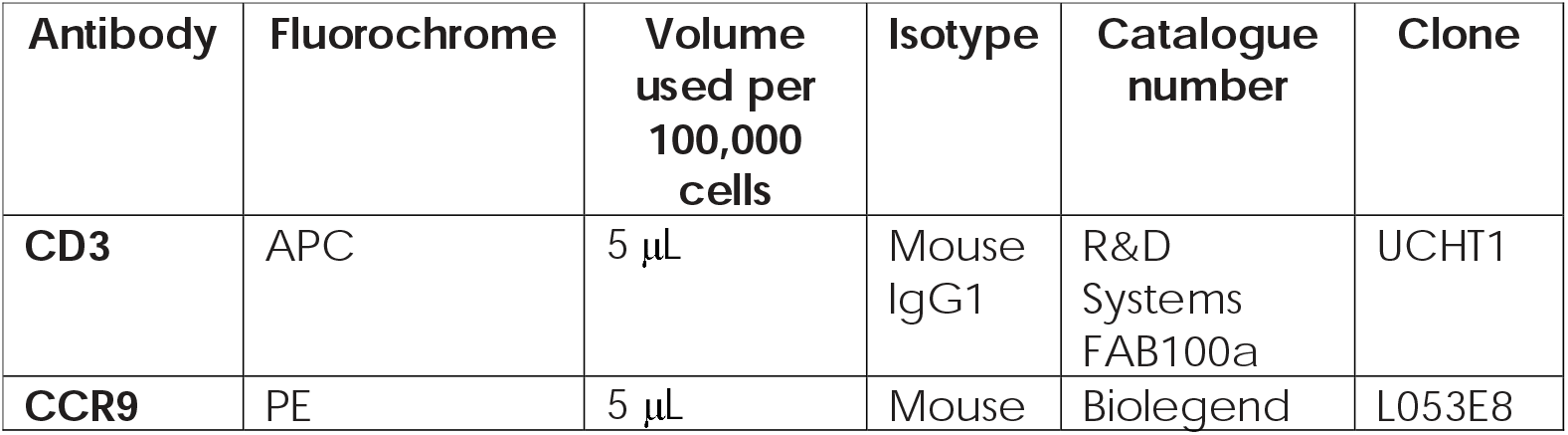
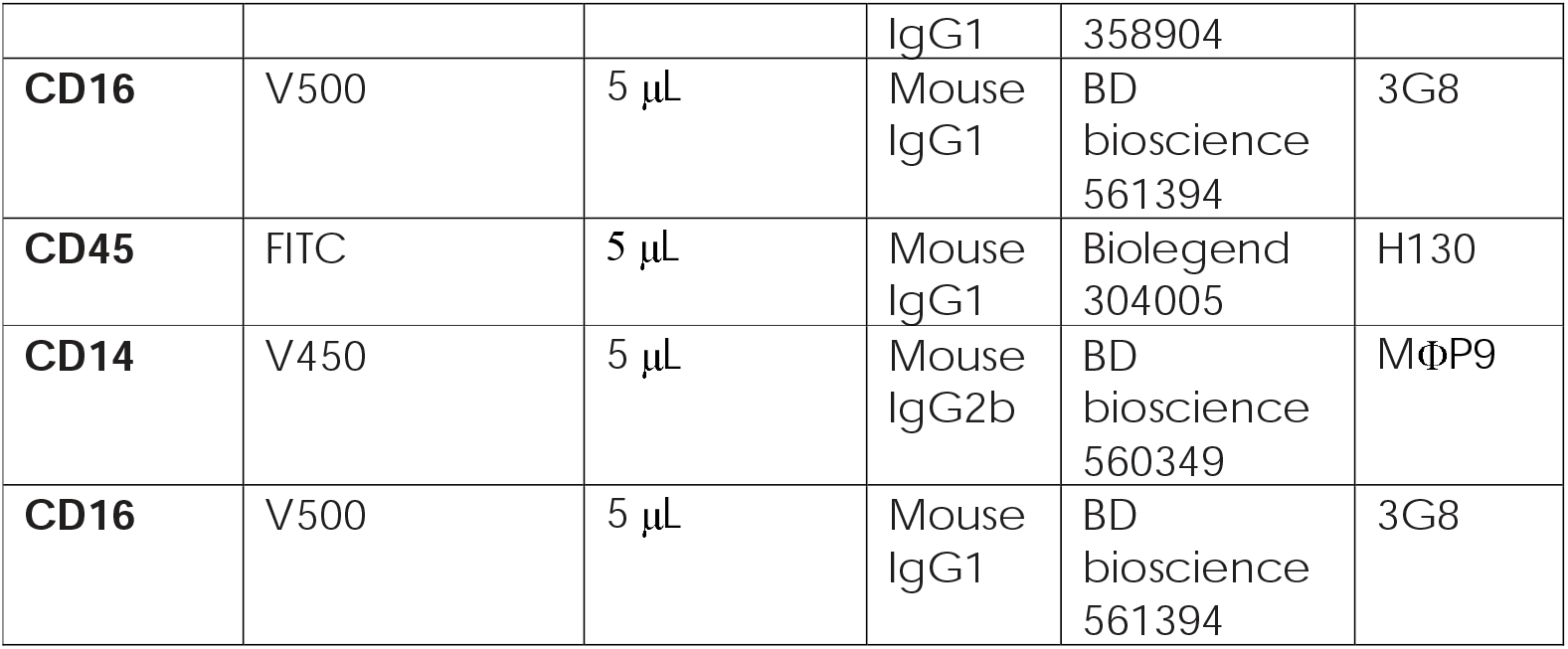

### Flow Based Adhesion Assay

Slides with six microchannels (Ibidi, Germany) were coated with rat-tail collagen (RTC) diluted at 1:100 in sterile water, by adding RTC to each channel for thirty minutes and then removing the remainder. Isolated HSEC were suspended at 1× 10^6^ml, cultured in the channels at 37 ^°^C/5% CO_2_ until confluent, then stimulated with TNFα and IFNγ (Peprotech, New Jersey, USA, both10ng/mL) for 24 hours.

Whole blood was mixed with an equal volume of PBS. Thirty mL of this mixture was layered over 20 mL of Lympholyte (CedarLane Labs, Canada) and centrifuged at 2000rpm/870g for 25 minutes. The resulting mononuclear cell layer was removed and washed a further three times suspending in RPMI and centrigfuing at 2000rpm for 5 minutes. Leukocytes were used immediately.

RPMI with a variety of concentrations of CCL25 was flowed over the HSEC at physiological levels of shear stress of (0.28ml/minute) for 5 minutes, then RPMI without CCL25 was flowed for a further 2 minutes. Suspended leukocytes (0.5×10^6^ cells/mL) in RPMI were then flowed over the HSEC for 5 minutes. Images were recorded for the final two minutes of this period. After 5 minutes, media without lymphocytes was flowed at the same rate, and then further images recorded for another 2 minutes.

Images were reviewed and analysed with regard to numbers of cells which were rolling, static, shape changing or transmigrated. Numbers of cells counted were normalised to number of cells/mm2/cells infused using the formula (number cells / (flow * bolus * area)) * (1 / # million cells) (Lalor *et al*., 1997).

### Statistical Analysis

Averages are expressed as mean and standard error of mean (SEM) for normally expressed data, and median and interquartile range (IQR) for skewed data. Normality was assessed with the Kolmogorov-Smirnov test. Normally distributed data were compared between groups with student’s t-test, and the Mann-Whitney test used for skewed data. Variance across multiple groups, for example over a range of concentrations was analysed with one-way analysis of variance (ANOVA). Data were analysed using Prism version 5 (California, USA).

## Results

The concentration of CCL25 in human serum measured by enzyme-linked immunosorbent assay (ELISA) correlated with body mass index (BMI) (Pearson r^2^ 0.500, p<0.001) (**figure 1A**). When considering obesity as a binary variable, i.e. non-obese vs. obese, a significant difference in mean serum CCL25 concentration was observed (median 0.624 pg/mL, IQR 1.10 vs. 2.894 pg/mL, IQR 1.233, Mann Whitney test p<0.001)(**supplementary figure 1A**). Image analysis of immunohistochemical staining for CCL25 in adipose tissue demonstrated elevated chemokine levels in obesity (one-way ANOVA p=0.04) (**figure 1B, supplementary figure 2**) and gene expression analysis detected *CCL25* transcripts in AT at much greater levels than that found in liver tissue (**figure 1C**). Hepatic gene expression of *CCL25* increased across the spectrum of NAFLD, with highest expression in histologically confirmed non-alcoholic steatohepatitis (NASH). In contrast to gene expression, the quantity of CCL25 protein was comparable between AT and liver, at least in the more advanced stages of NAFLD (**figure 1D**).

**Figure 1:**
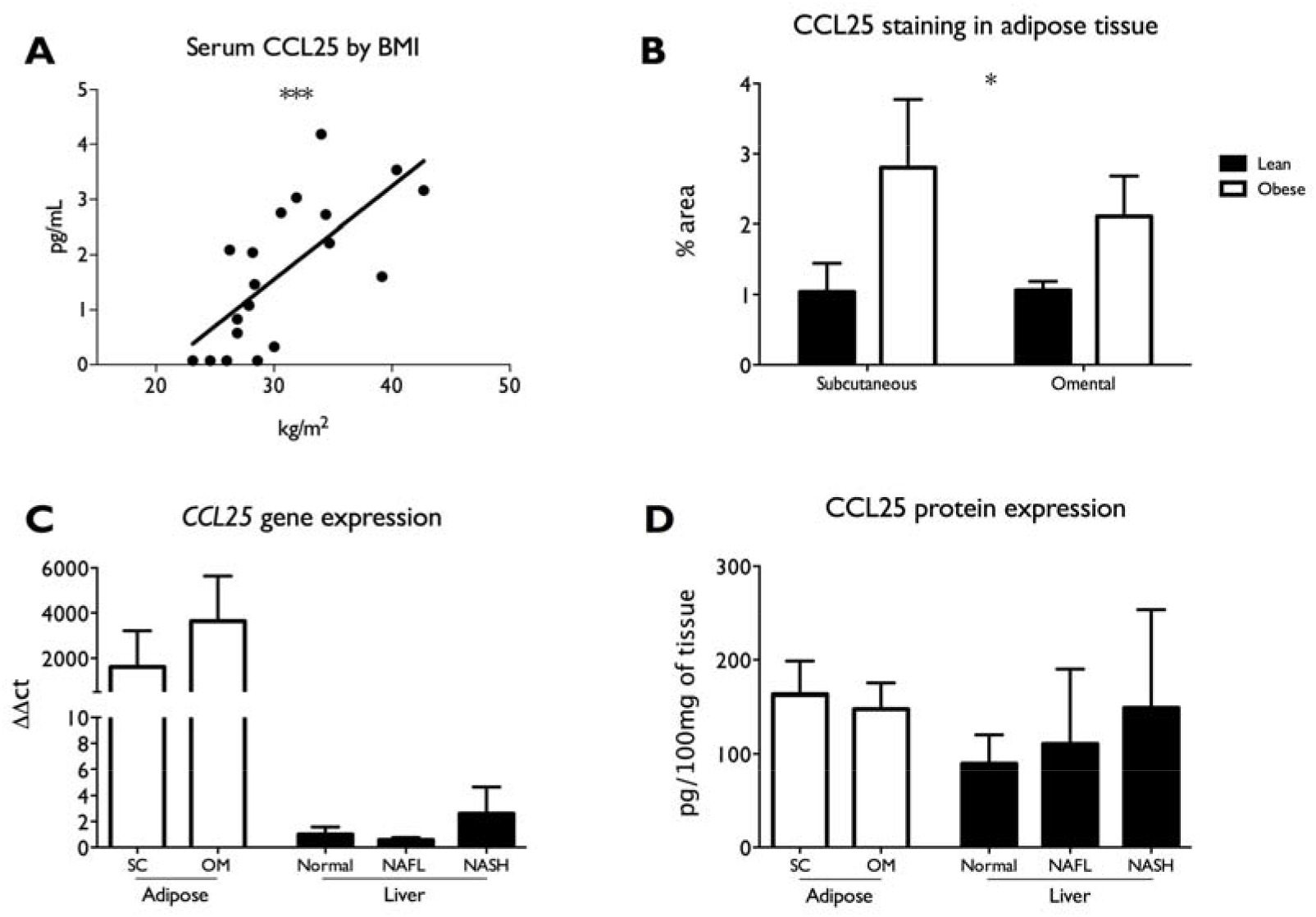
A serum concentration of CCL25 measured by ELISA showed a positive correlation with BMI (Pearson r ^2^ 0.500, p<0.001). B measurement of protein levels of CCL25 measured by immunohistochemical staining showed an increase in CCL25 levels in obesity (* p<0.05 by one-way ANOVA). C Gene expression of CCL25 in adipose and liver tissue measured by rt-PCR showed greater expression in adipose tissue. D Protein levels of CCL25 were comparable between adipose and liver tissue.

**Figure 2.**
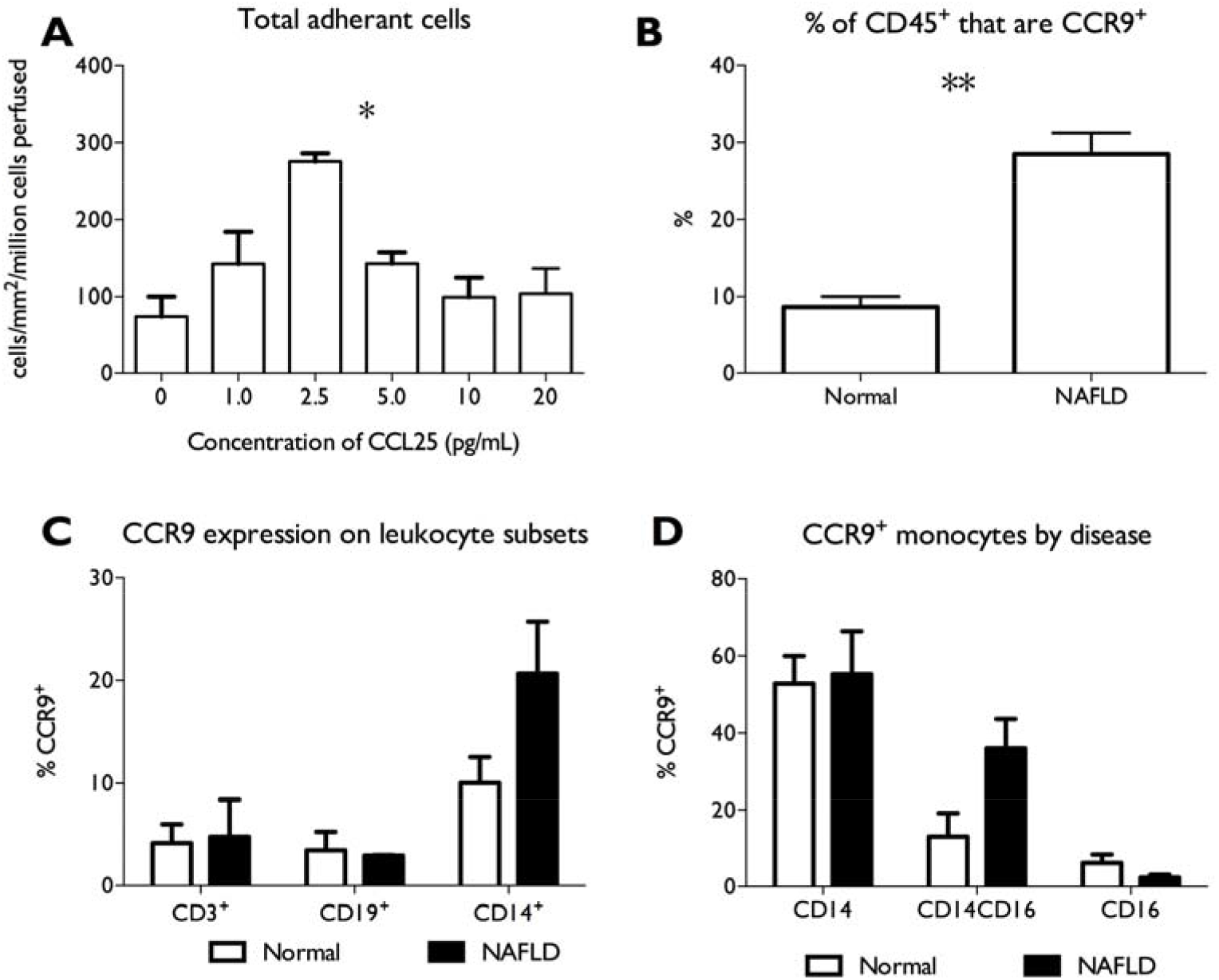
**A** adherence of leukocytes to HSEC after pre-treatment with CCL25 (* one-way ANOVA p<0.05) **B** Total CCR9^+^ cells isolated from normal and NAFLD liver tissue (** student’s t-test p<0.01). **C** CCR9 expression on leukocyte subsets from normal and NAFLD liver tissue **D** CCR9 expression on liver-infiltrating monocytes in normal and NAFLD liver tissue

We postulated that increased secretion of CCL25 from AT in obesity influences hepatic inflammation, and contributes to the pool of hepatic CCL25. We used a well described flow-based adhesion model of the hepatic sinusoid, with primary human hepatic sinusoidal endothelial cells (HSEC) and peripheral blood leukocytes, to determine whether exogenous CCL25 promotes leukocyte trafficking ^[11]^. Media supplemented with varying concentrations of recombinant soluble CCL25 was passed over a monolayer of HSEC that had been pre-stimulated with pro-inflammatory cytokines, and then isolated leukocytes were flowed over the HSEC at a physiological flow rate of 0.05 Pa. When interactions between leukocytes and HSEC were directly visualized and quantified, a clear effect of pre-treatment with CCL25 was observed where leukocyte trafficking through HSEC varied with concentration of CCL25, with a classical ‘bell-shaped’ effect commonly observed with chemokines, peaking at 2.5 pg/ mL (**figure 2A**), that influenced all stages of the leukocyte adhesion cascade: adhesion, activation and transmigration (**supplementary figure 2**). The peak effect was observed at a concentration consistent with the mean concentration observed in serum from obese individuals (**supplementary figure 1A**).

The cognate receptor for CCL25 is CC-chemokine receptor 9 (CCR9). Using flow cytometry, we analyzed the presence and type of CCR9^+^ leukocytes in liver tissue from patients with NAFLD and pathological control tissue. The proportion of CD45^+^CCR9^+^ cells was elevated in liver tissue from individuals with NAFLD when compared to control tissue (25.8% vs 8.7%, students t-test p=0.003)(**figure 2B**). CCR9 was predominantly expressed by CD14^+^ monocytes isolated from both pathologically normal and diseased liver tissue (**figure 2C**). Analysis of monocyte subsets showed that while the proportion of CCR9^+^ cells was greatest in ‘classical’ CD14^+^CD16^-^ monocytes, NAFLD was associated with an increase in the proportion of CD14^+^CD16^+^ monocytes expressing CCR9 (**figure 2D**). This subset of tissue infiltrating monocytes is associated with inflammation and fibrosis ^[12]^.

## Conclusions

NAFLD is almost always accompanied by obesity, and obesity is often a cofactor in the progression of other chronic liver disease, most notably alcohol related liver disease ^[13]^ but also viral hepatitis ^[14; 15]^. These data describe a novel mechanism of inflammatory cross-talk between adipose and hepatic tissue through the expression of CCL25 from adipose tissue and subsequent effects on hepatic immune cell trafficking.

CCL25 has been reported as playing a pro-inflammatory role in other inflammatory gastroenterological and hepatic disease. Indeed, CCR9 antagonism has been considered as a potential treatment for inflammatory bowel disease. These data are the first to identify a role for CCL25 in NAFLD. We propose that in obesity, over-expression of CCL25 in adipose tissue and subsequent release into the circulation promotes immune cell recruitment in the liver and an increased prevalence of CCR9^+^ intrahepatic CD14^+^CD16^+^ monocytes. Previous work from our group has shown that this intermediate group of monocytes is found in increased frequency in fatty liver disease and is associated with tissue damage and scarring ^[12]^. In a common disease such as NAFLD, that does not yet have accepted treatment options, this represents an important addition to our understanding of the disease and a pathway that might be amenable to therapy.

## Data Availability

Data are not publicly available but can sharing of anonymised experimental data can be discussed with the corresponding author

---

**Supplementary figure 1:**
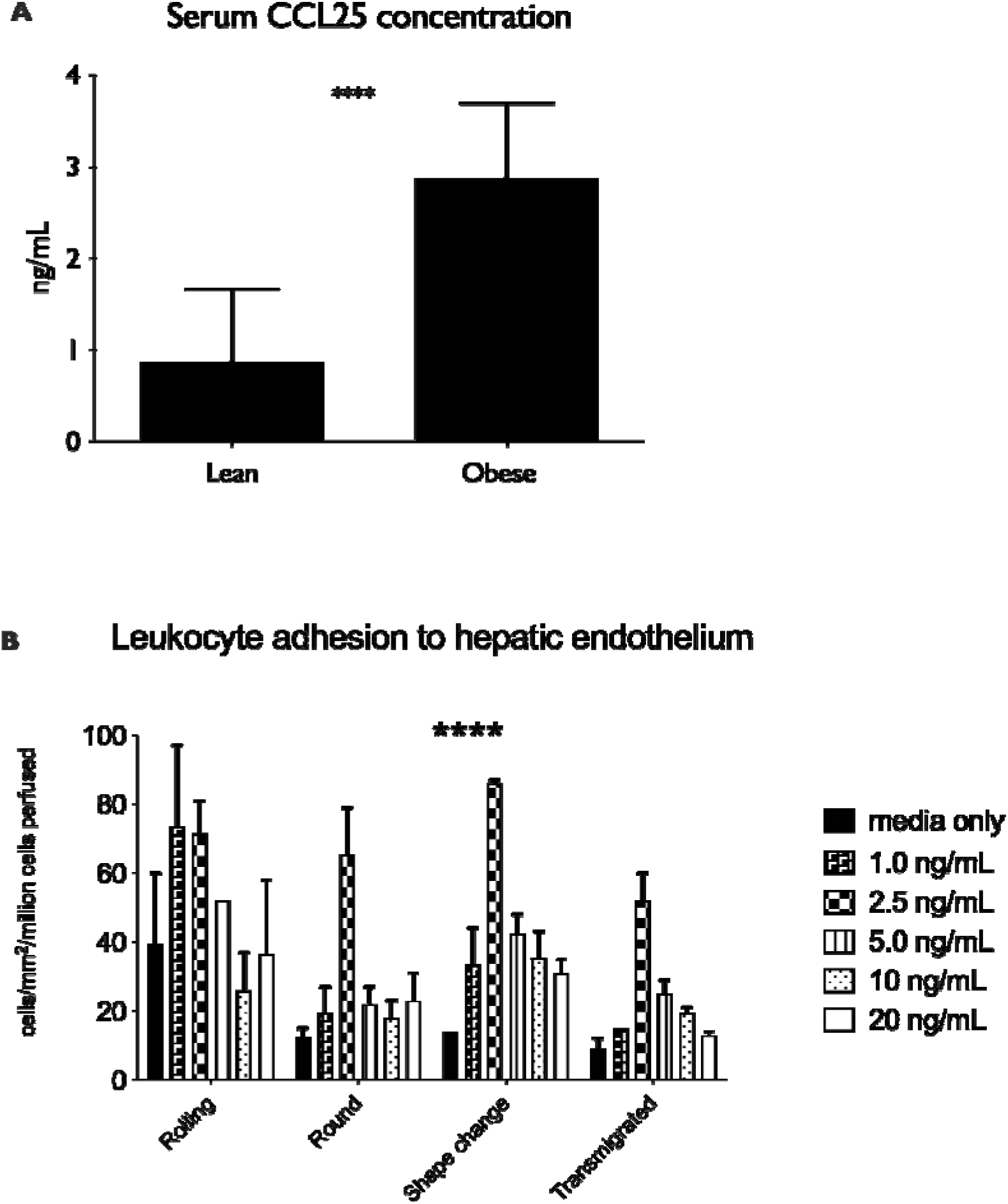
**A** serum concentration of CCL25 measured by ELISA in lean and obese individuals **** p<0.001 by Mann Whitney test **B** lymphocyte trafficking in response to pre-treatment of hepatic endothelial cells with recombinant CCL25 ****p<0.001 by two-way ANOVA

